# The impact of hypertension on the dose-response association between physical activity and stroke: A cohort study among 139,930 adults from the Netherlands

**DOI:** 10.1101/2023.11.16.23298659

**Authors:** Hannah L McLellan, Ellen A Dawson, Thijs MH Eijsvogels, Dick HJ Thijssen, Esmée A Bakker

## Abstract

**Background:** There is a strong dose-response relationship between regular physical activity (PA) and stroke risk. However, this relationship is attenuated in the presence of cardiovascular risk. This study aimed to compare the dose-response relationship between PA and stroke between normo- and hypertensive individuals.

**Methods:** A cohort study including 139,930 individuals was performed (median follow up: 6.75 years). Participants were stratified at baseline as hypertensive or normotensive and were categorised into quartiles of lowest (Q1) to highest (Q4) moderate-to-vigorous (MV), self-reported PA. Primary outcome was stroke. Cox regression was used to estimate hazard ratios (HRs) and 95% confidence intervals. The main analyses were stratified on baseline blood pressure and adjusted for confounders. Additionally, hypertensives were stratified into medicated or not medicated.

**Results:** Compared to Q1, adjusted HRs were 0.87 (0.69-1.10, *P*=0.23) for Q2, 0.75 (0.59-0.95, *P*=0.02) for Q3, and 0.94 (0.74-1.20, *P*=0.64) for Q4 in the total population. In the stratified analyses, HRs for individuals with normotension were 0.79 (0.50-1.25, *P*=0.32), 0.75 (0.48-1.18, *P*=0.22), 0.97 (0.62-1.51, *P*=0.90) for MVPA Q2 to Q4, respectively. In hypertensive individuals, HRs compared to Q1 were 0.89 (0.68-1.17, *P*=0.41), 0.74 (0.56-0.98, *P*=0.03), 0.92 (0.69-1.23, *P*=0.56) for Q2-Q4, respectively. There was no significant interaction between PA and hypertensive status. A smaller benefit of MVPA in medicated hypertensives compared to non-medicated was observed, however the dose-response association was similar.

**Conclusion:** MVPA reduces stroke risk in the total population (Q3), which is not affected by the presence of hypertension. Use of anti-hypertensive medication may interfere with the impact of MVPA on stroke risk.

**Novelty and Relevance:** *What is new?:* Presence of cardiovascular disease or cardiovascular disease risk factors may alter the dose-response relationship between regular physical activity and major adverse cardiovascular events (MACE) and mortality. No previous study has investigated whether hypertension *per se* alters the dose-response relationship between regular moderate-to-vigorous physical activity (MVPA) and stroke risk.

*What is relevant?:* This large cohort study demonstrates the importance of regular MVPA on stroke risk reduction particularly in individuals with hypertension. Furthermore, this study provides preliminary indication that the use of antihypertensive medication may alter this association.

*Clinical/Pathophysiological implications?:* The presence of hypertension does not alter the dose-response association between MVPA and stroke. Importantly, this highlights that MVPA is beneficial in reducing the risk of stroke, even in individuals diagnosed with hypertension. Although this cohort study cannot make solid conclusions on the influence of antihypertensive medication, this study reinforces the importance of regular physical activity on the clinical outcome of stroke.

## Introduction

Stroke remains a significant health problem, with a predicted increased incidence of 34% between 2015 and 2035^1^. This highlights the need for preventive strategies to lower the risk for stroke, and there by reducing the health and socio-economic effects pertaining to stroke.

Several studies found potential health benefits of physical activity (PA) in the prevention of stroke ^2–4^. For example, ≥30 minutes of moderate-to-vigorous (MV) PA during 1 to 2 times per week resulted in a 16% risk reduction for the risk of stroke compared to inactive individuals^3^. Similarly, 3 to 4 times/week and ≥5 times/week of MVPA sessions were also associated with 21 and 22% a lower risk of stroke compared to inactivity, respectively^3^. In addition to the risk of stroke, MVPA is also associated with a lower risk of developing hypertension, an important modifiable risk factors for stroke ^5^. Some studies have suggested that the positive responses to PA may become stronger with the prevalence of risk factors, such as hypertension^6–10^. Supporting this, a recent study in 142,493 adults found that individuals with cardiovascular risk factors may have larger benefits of MVPA^11^, as the dose-response association between MVPA and major cardiovascular events and mortality became stronger and contained larger risk reductions in individuals with risk factors compared to individuals without risk factors or with established CVD. The dose-response relation between physical activity and the risk for stroke, therefore, may be affected by presence of hypertension, one of the key risk factors. Furthermore, anti-hypertensive pharmacological treatments may also interfere with the benefits of exercise training on the risk for clinical events.

Therefore, the aim of our study was to examine the dose-response association between MVPA and stroke and to determine whether this association differed between those with *a priori* hypertension and normotension. Secondly, we aimed to evaluate whether the use of anti-hypertensive medication alters this dose-response association. In line with recent work, we expect that those with hypertension will demonstrate a stronger inverse association between MVPA and the risk of stroke. In addition, we hypothesize that anti-hypertensive medication may attenuate the benefits of MVPA on stroke, as recent exercise training studies evaluating the impact of drugs on exercise mediated cardiovascular adaptations was blunted with simvastatin^12^ and angiotensin converting enzyme inhibitors whilst angiotensin-converting^13^.

## Methods

### Study population

This study used data from the Lifeline Cohort Study. Lifelines is a multi-disciplinary prospective population-based cohort study examining in a unique three-generation design the health and health-related behaviours of 167,729 persons living in the North of the Netherlands. It employs a broad range of investigative procedures in assessing the biomedical, socio-demographic, behavioural, physical and psychological factors which contribute to the health and disease of the general population, with a special focus on multi-morbidity and complex genetics^13,14^. All individuals who lived in the northern Netherlands were eligible to take part, other than those with 1) severe psychiatric or physical illness (e.g. individuals with cancer and associated reduced life expectancy), 2) life expectancy <5 years; and 3) lack of fluency in Dutch. Participants who were ≥18 years old were include (N=152,737). Participants were excluded (N=12,807) due to 1) missing data for PA, blood pressure and CVD health status; 2) limited ability to be physically active; 3) inability to merge individual data with registry data; and 4) the presence of cardiovascular disease (**Supplemental Figure 1**). The Strengthening the Reporting of Observational Studies in Epidemiology (STROBE) guideline was used to report our findings^15^. All participants provided written informed consent. The Lifelines Cohort study was conducted according to the principles of the Declaration of Helsinki and approved by the University Medical Centre Groningen Medical Ethical Committee.

### Measurements

#### Physical examination and questionnaire

Participants visited one of the Lifeline research sites for a physical examination and completed a baseline questionnaire between 2006 and 2013. Baseline data was collected for 167,729 participants, aged from 18 to 93 years. Every 1.5 years a follow up questionnaire was administered to assess the occurrence of a stroke.

The physical examination included anthropometric assessment, blood pressure (BP) measurements and blood draw. Anthropometrics included height (Stadiometer) and weight (Standard weighing scale), which were used to calculate body mass index (BMI; kg/m^2^). Ten blood pressure measurements were taken during a 10-minute period using an automated sphygmomanometer (Dynamap, PRO 100 or PRO 100V2) placed around the upper right arm (or the left arm if contraindications were present) with participants in a seated position, the two last successive blood pressures were averaged. Blood samples were taken after >8 hour fasting for measurements of low-density lipoprotein (LDL), glucose and serum creatinine. Renal function (estimated glomerular filtration rate, eGFR) was estimated.^16^

Baseline questionnaires included questions on demographics, health status and lifestyle. Demographics included age, sex, postal code, income and education level. Income was estimated using the postal codes and data of Statistics Netherlands^17^ when not reported. Education level was categorised in low, moderate and high. Medical history included self-reported information on medication use, presence of CVD, comorbidities and other illnesses including cancer, arthritis, multiple sclerosis and Parkinson disease. Lifestyle factors included smoking status, alcohol consumption, and hours of sleep per night. Smoking status was categorised as currently, previously, and never. High alcohol consumption was defined as >14 drinks/week or >4 drinks/day for men and >7 drink/week or > 3 drink/day for women^18^.

#### Habitual Physical Activity Volumes

Baseline physical activity was assessed using the Short Questionnaire to Assess Health-enhancing Physical Activity (SQUASH). SQUASH divides habitual physical activity into transportation, occupation, household and leisure domains, and asks for the duration and intensity of an individual’s typical weekly activities over the past 3 months^19^. Weekly physical activities were converted to the average amount of metabolic equivalent of task (MET) minutes per week based^20^. MET minutes were calculated by multiplying the MET values of each activity by the duration (i.e., minutes/week). Only activities with ≥3 MET value were included, since these activities relate to moderate to vigorous activities as specified in the World Health Organisation PA guidelines^21^. Individuals were categorised into 4 quartiles of least (Q1) to most (Q4) physically active based on self-reported MVPA volumes. Q1 included the least active individuals who spent <1830 MET min/week, Q2 included individuals with 1830-3617 MET min/week, Q3 included individuals with 3618-7175 MET min/week and Q4 included individuals with >7175 MET min/week.

#### Clinical outcomes

The primary endpoint was the occurrence of a stroke. Hospital registry data of statistics Netherlands were used to determine the primary outcome. When hospital registry data were not available, self-reported stroke during follow-up were used instead. Self-reported stroke was measures after a median follow-up of 1.1, 2.1, and 3.8 years. The date at which the questionnaire was completed, was used for the event date of self-reported stroke. Participants were followed until the first stroke event. Participants who did not reach the endpoint were censored at the end of the last assessment or at the date they died, whichever occurred first.

### Hypertension

#### Hypertension

Participants were divided as hypertensive or normotensive based on the BP evaluation performed at baseline, whilst using the recent classifications of hypertension following the updated National Institute for Health Care Excellence (NICE) guidelines (i.e. ≥130/80 mmHg). In addition, those who reported being diagnosed by a physician with hypertension and used blood pressure lowering medication were included in the hypertensive group. Those who did not report being diagnosed with hypertension and had a baseline blood pressure of ≤129/79 mmHg were categorised into the normotensive group.

#### Medicated

For our secondary analysis, we have divided the hypertensive group into a medicated and non-medicated group. Individuals were categorised into the medicated group if they were prescribed Angiotensin Converting Enzyme (ACE) inhibitors, angiotensin blockers, beta blockers, diuretics, or calcium antagonists.

### Statistical analyses

Baseline characteristics were described for the normo- and hypertensive group. Normally distributed data were presented with mean (± standard deviation; SD) and non-normally distributed data were presented with the median (interquartile range; Q_25_ to Q_75_).

Stratified Kaplan–Meier curves and log-rank tests were performed to assess differences in the outcome between the quartiles of physical activity. Multivariable Cox regression model was conducted to estimate the association between physical activity and stroke using hazard ratios (HRs) with 95% confidence intervals (CIs). First, an unadjusted model was performed (model 1). Model 2 was adjusted for age (years), sex (male/female), body mass index (BMI; (kg/m^2^)), income (per 1,000 euros), education level (low/moderate/high), smoking (pack years), kidney function (glomerular filtration rate, mL/min/1.73m^2^), glucose (mmol/L) and low-density lipo-protein (LDL; mmol/L). Model 3 included the variables of model 2 and was additionally adjusted for systolic blood pressure (mmHg), diastolic blood pressure (mmHg), use of acetylsalicyclic acid, anti-platelets and anti-hypertensive medication. The analyses were performed for the total population and were stratified for hypertensive and normotensive individuals. We also tested interaction in terms of blood pressure (normotensive versus hypertensive) and the 4 quartiles of MVPA. In addition, to examine whether medication use altered the dose-response association of MVPA in hypertensive individuals, we performed stratified analyses and tested the interaction term.

Since the number of missing values was relatively small (<14%), the complete case was used for the analyses. All statistical analyses were performed in R version 3.5.2. P values < 0.05 were considered statistically significant.

## Results

### Study population

In total, 139,930 participants were included in our analysis. 41% of the population was male with an overall mean age of 41 (13) years (**Table 1**). The median follow up was 6.75 years [Q_25_ 5.83; Q_75_ 7.92] with a total number of 640 strokes (0.46%; **Figure 1**). Individuals with hypertension were more often male, older, had a lower education level and higher BMI, used more often medication and had more comorbidities. Within the hypertensive group, medicated individuals were more often female, older, had a lower education level, and had more comorbidities compared to non-medicated (**Supplemental table 1**).

**Figure 1:**
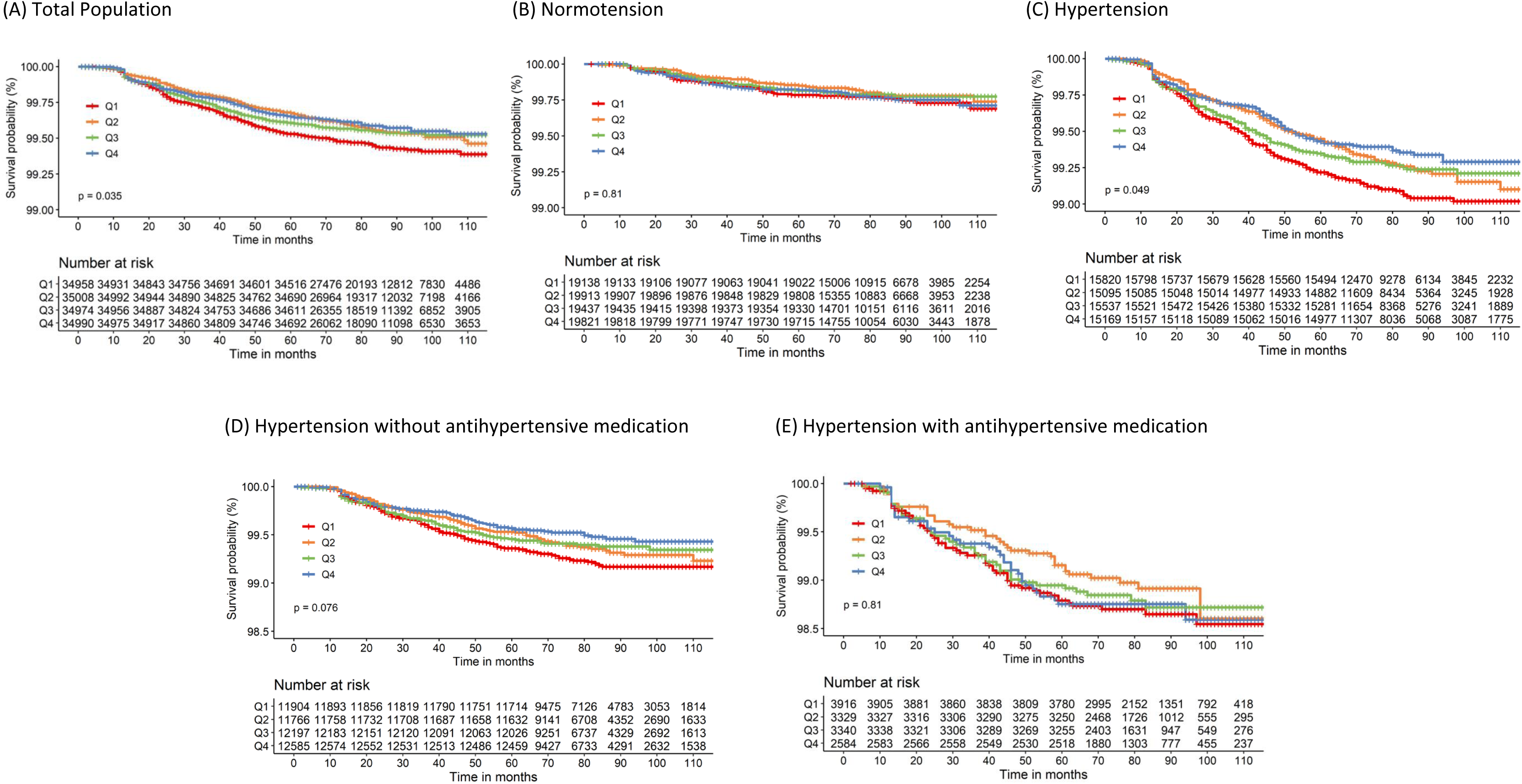
Unadjusted Kaplan–Meier estimates of stroke occurrence for quartiles (Q) of moderate-to-vigorous physical activity during follow-up stratified for total population(A), individuals with normotension (B), and individuals with hypertension (C), individuals with hypertension who are not taking anti-hypertensive medication (D) and individuals with hypertension who are taking antihypertensive medication.

**Table 1.**
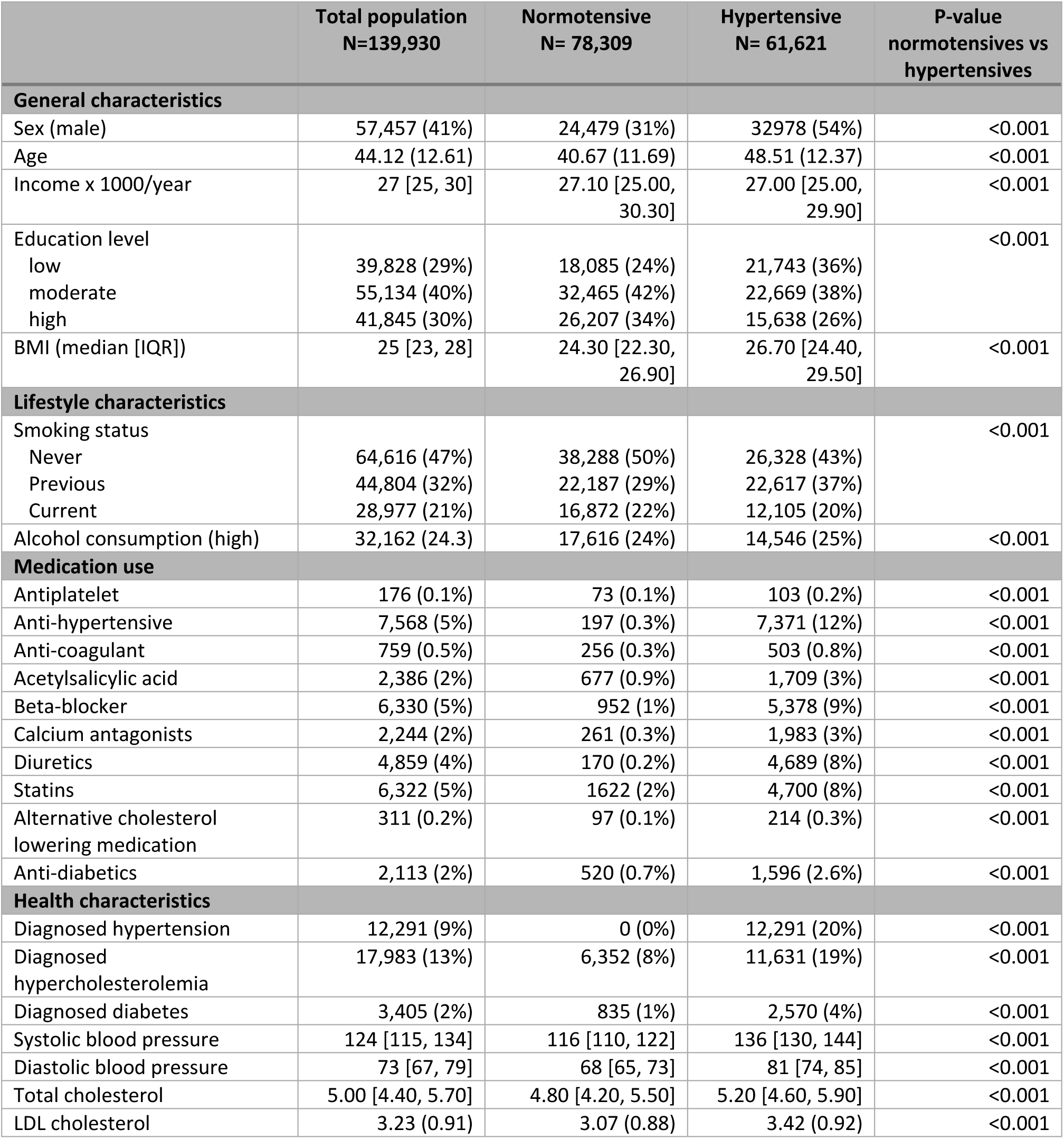
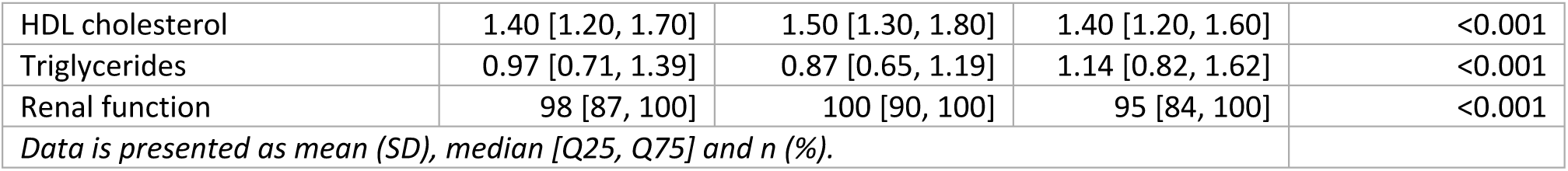
Baseline characteristics for the total population (n=139,949) and individuals with normotension (N=78,309) and hypertension (n=61,621). Data are presented as mean (SD), median [Q25,Q75], and *n* (%).

### Physical activity and the risk of stroke

There was a significant negative and linear trend between MVPA and stroke risk for the total population and in the hypertensive subgroup, which disappeared partly after adjusting for confounding factors (Model 2 and 3; **Table 2**). After adjustments for confounders (model 3) and using MVPA categories, a significantly lower HR was found in Q3 compared to the least active individuals (Q1) within the total population and in individuals with hypertension (**Table 2 and** **Figure 2**). No significant association was found in Q2 and Q4 compared to Q1. Although we found no significant association between MVPA and stroke risk in normotensive individuals, estimated HRs were largely comparable to the hypertensive participants. In addition, there was no significant interaction between each of the MVPA quartiles for stroke risk and normotension versus hypertension (P>0.05), suggesting that dose-response curves between MVPA and stroke are not different between normotensive and hypertensive individuals.

**Figure 2.**
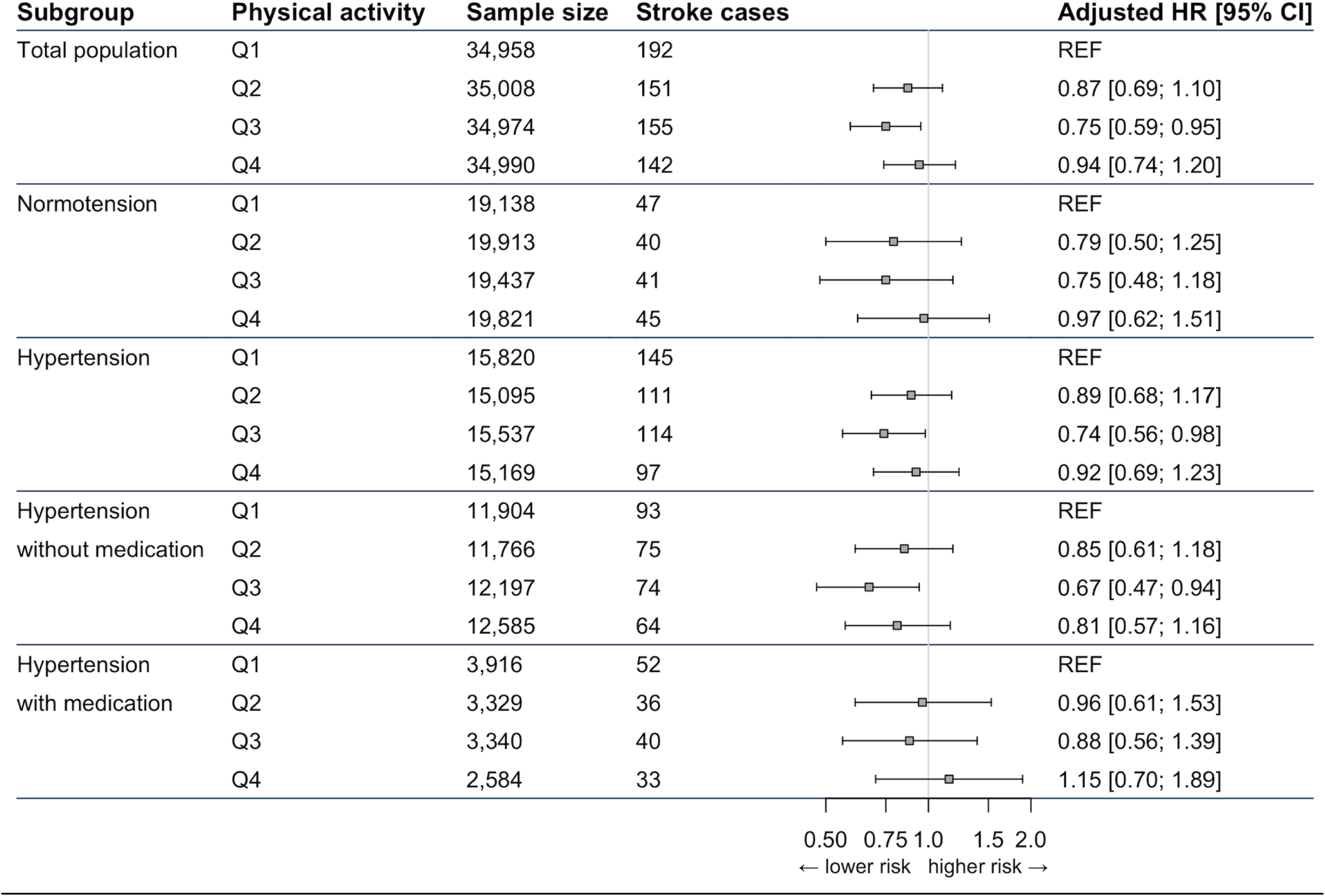
Forrest plot of the quartiles (Q) of MVPA associated with stroke risk for the total population and stratified for blood pressure and medication use. HRs were adjusted for age, sex, BMI, income, education level, alcohol, smoking (pack years), kidney function, BMI, serum glucose, systolic blood pressure, diastolic blood pressure, low-density lipoprotein cholesterol., use of acetylsalicylic acid, anti-platelets, antihypertensive medication (model 3). 95% CI, 95% confidence interval; HR, hazard ratio.

**Table 2.**
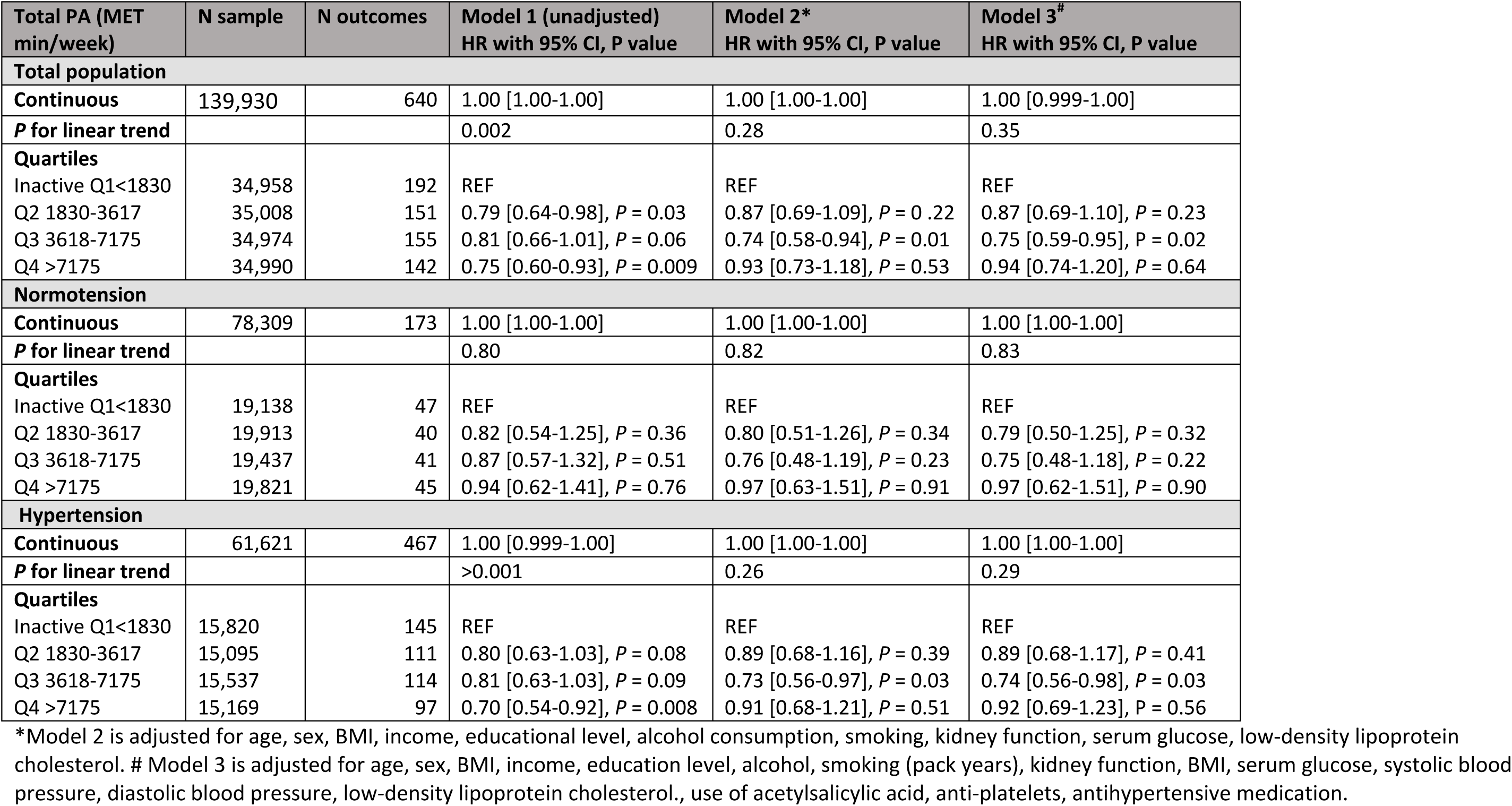
Hazard ratios (HR) with 95% confidence intervals (95% CI) for the association between moderate-to-vigorous physical activity and stroke.

### Physical activity and stroke risk in hypertensives: impact of medication use

Within hypertensive individuals, after adjustment for confounders, there was a significant reduction Q3 compared to Q1 for MVPA and stroke risk in those without medication (**Supplemental 2** and **Figure 2**). In medicated hypertensive individuals, no significant association was found between PA and stroke risk, with estimated HRs being slightly higher compared to the non-medicated hypertensive population (**Supplemental 2** and **Figure 2**). However, the p-value for interaction between PA and medication use was non-significant (P>0.05).

## Discussion

Previous research has demonstrated the importance of PA on stroke risk reduction, however emerging research has suggested that the dose-dependent association of regular PA may be altered in the presence of cardiovascular risk factors and/or medication use therefore we aimed to further investigate this. This study presents the following findings. First, we reinforce the benefits of regular PA, as engagement in MVPA is associated with a lower risk for stroke, which remained significant upon adjustment for potential confounders. Second, the shape of the dose-response association does not importantly differ between hypertensive (n=61,621) and normotensive (n=78,309) individuals. However, the association remained only significant in the hypertensives after stratification. Thirdly, the benefits of regular MVPA seem to be only stronger in non-medicated individuals with hypertension, but further studies are necessary to confirm this preliminary result. Our findings highlight that a physically active lifestyle is beneficial for reducing stroke risk.

Our study reinforces findings from previous studies highlighting that more PA is associated with a reduction in the risk for stroke in the general population. For example, a study in the USA, with a median follow up of 18.8 years and 648 ischemic stroke events, showed a significant reduction in ischemic stroke (and other subtypes) with increasing PA levels among middle-aged men and women. Another prospective cohort study in South Koreans with 13 years follow-up showed that exercising between 3–4 or 5–6 times/week showed the lowest stroke risk compared to physically inactive individuals^22^. In contrast to others, our study further examined whether the association between MVPA and stroke depends on BP levels. We found that the shape of the dose-response association between MVPA and stroke risk was not significantly different between individuals with normotension or hypertension. Nevertheless, the association was not significant in the normotensive group. A possible explanation for this observation is due to the low event rate present in the normotensive group (i.e., >60% less events in the normotensives *versus* the hypertensives). In addition, compared to other studies, our follow-up time was shorter (i.e., median 6.8 years) and we included a relatively young population, which suggests that a longer follow-up in the normotensives who have a lower *a priori* risk for stroke may be required. Furthermore, HRs in Q4 in the present study are higher that Q3, however this may be explained by possible measurement error in MVPA classification, Specifically, Q4 may contain individuals who exaggerated their PA levels but due to the nature of questionnaires and the absence of an objective PA measurement we can only speculate. Importantly, our findings show that the presence of hypertension does not attenuate the dose-response relationship between MVPA and stroke. This finding supports previous studies examining the effect of PA and cardiovascular disease and mortality ^11,23^. Our study provides an important public health message; that benefits from a physically active lifestyle in relation to stroke remain equally present in those who have already developed hypertension.

For our secondary aim, we compared the effects of PA on stroke risk between medicated and unmedicated hypertensive individuals. Whilst reinforcing our initial results, in that PA is associated with lower risks for stroke in unmedicated hypertensive individuals, such effects were not observed in medicated individuals with hypertension. One potential explanation of the latter observation is that the sample size and event rate of the medicated hypertensives was relatively low. Although the medicated hypertensive individuals might be underpowered, the analysis showed higher HRs in this subgroup, which might suggest an attenuated and smaller effect of MVPA on the risk of stroke in the medicated group of individuals with hypertension. One potential explanation could be that those within the medicated group had higher BP levels and/or disease severity. Previous research has suggested that those with established CVD may not benefit to the same extent as healthy individuals ^9–11^. In addition, the attenuated effect may relate to the use of medication itself. However, previous work is conflicting whether antihypertensive medication may interfere or potentiate the benefits of PA or exercise training ^24–27^. Apart from the antihypertensive medication, other drugs that are commonly prescribed to hypertensive individuals such as statins (21% in our medicated hypertensives) may also have an effect on the benefits of PA^12^. Therefore, further research is warranted to understand whether antihypertensive medication affects the benefits of PA on stroke risk. Nonetheless, we believe it is important to emphasise that, even in the presence of hypertension and medication use, other research has suggested that when combined, exercise and medication strengthen the BP effects of medication alone and should be recommended and implemented as indicated according to existent professional treatment algorithms ^28,29^.

## Strengths and limitations

A particular strength of this study is that it includes a large population (n = 139,930) with outcome data based on national hospital registry data. One potential limitation of our study is that MVPA was self-reported, which is susceptible for overestimating true PA volumes^30^. Nonetheless, categorising individuals across quartiles allowed comparison between the least active individuals and those engaged in larger volumes of MVPA, and ultimately self-reported MVPA data is likely to underestimate the true effect of MVPA on health benefits^30^. Another limitation relates to the relatively low event rate of stroke in the normotensive population, and relatively small sample size of medicated hypertensive individuals. Nonetheless, HRs in both populations, albeit with larger confidence intervals, provide insight into the effects of PA.

## Perspectives

Our study reinforces the benefits of regular MVPA on stroke risk reduction, especially in hypertensive individuals. However, we found that the shape of dose-dependent association between MVPA and risk of stroke was not different between individuals with hypertension and normotension. Furthermore, our data provide preliminary support that in individuals with hypertension, the use of antihypertensive medication may be associated with a smaller benefit of MVPA on the risk of stroke, albeit effects of PA in this subgroup have unlikely disappeared. Future studies are warranted to better understand the effects of regular MVPA on the risk of stroke in medicated hypertensive individuals and the potential underlying mechanisms, which can aid in optimising the benefits of MVPA in the prevention of stroke.

## Data Availability

The data can only be accessed by Statistics Netherlands

https://www.lifelines.nl/researcher/cohort-and-biobank

## Acknowledgments

The authors wish to acknowledge the services of the Lifelines Cohort Study, the contributing research centres delivering data to Lifelines, and all the study participants.

## Nonstandard Abbreviations and Acronyms

PA: Physical activity
MVPA: Moderate-to-vigorous physical activity

---

**Supplemental Figure 1.**
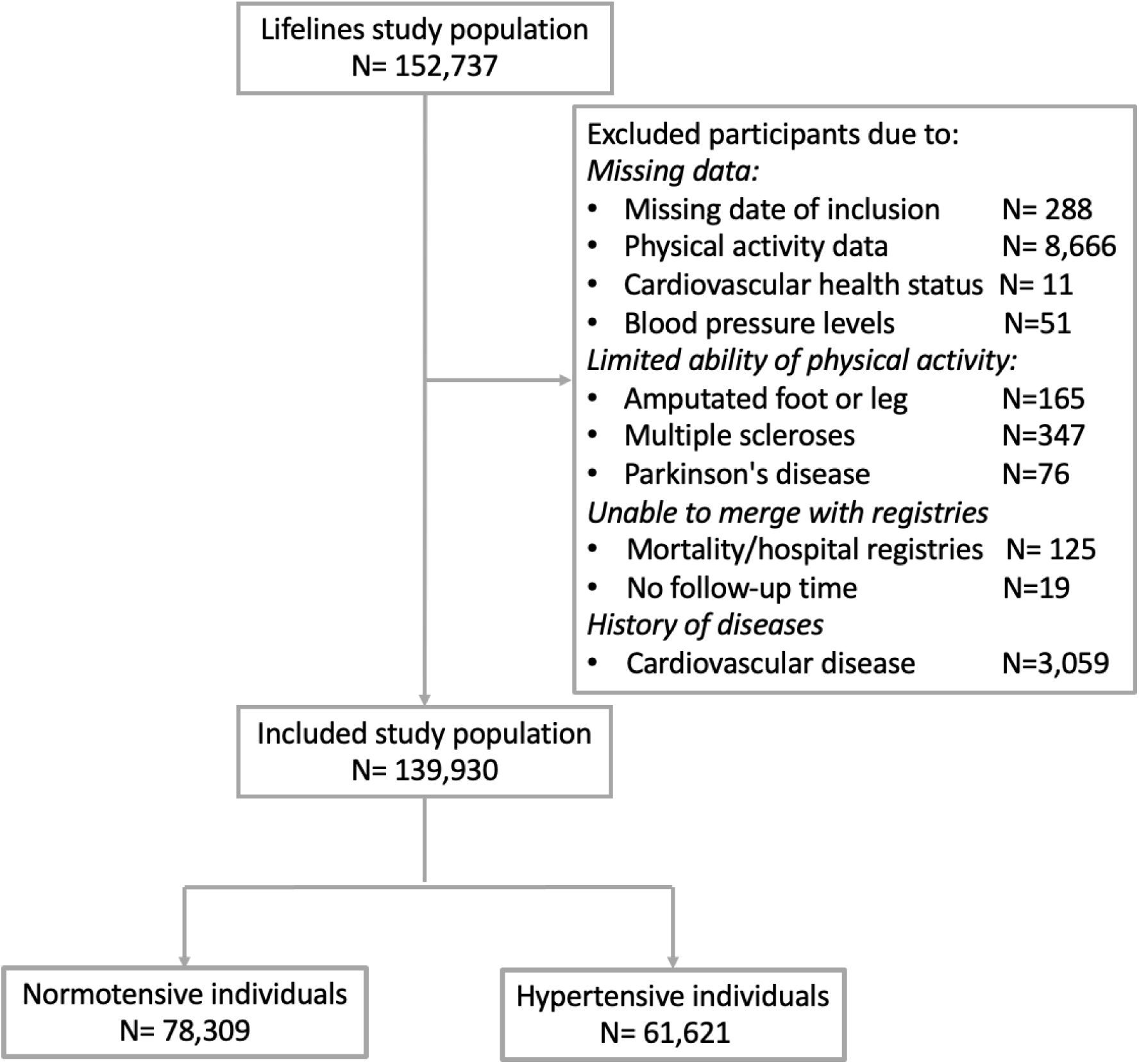
Flow chart of included study population.

**Supplemental table 1.** Baseline table of individuals with hypertension.

| General characteristics | Hypertensives without medication<br>(N=48,452) | Hypertensives with medication<br>(N=13,169) | p-value |
| --- | --- | --- | --- |
| Sex (male) | 27,584 (57%) | 5,394 (41%) | <0.001 |
| Age | 46.43 (11.90) | 56.16 (10.95) | <0.001 |
| Income x 1000/year | 27 [25.00, 30.00] | 26.80 [24.90, 29.70] | <0.001 |
| Education level |  |  | <0.001 |
| low | 15797 (33%) | 5,946 (47%) |  |
| moderate | 18620 (39%) | 4,049 (32%) |  |
| high | 12943 (27%) | 2,695 (21%) |  |
| BMI (median [IQR]) | 26.40 [24.20, 29.10] | 27.90 [25.40, 31.10] | <0.001 |
| **Lifestyle characteristics** |  |  |  |
| Smoking status |  |  | <0.001 |
| Never | 21,246 (44%) | 5,082 (39%) |  |
| Previous | 16,474 (34%) | 6,143 (47%) |  |
| Current | 10,256 (21%) | 1,849 (14%) |  |
| Alcohol consumption (high) | 11,824 (26%) | 2,722 (21%) | <0.001 |
| **Medication use** |  |  |  |
| Antiplatelet | 47 (0.1%) | 56 (0.4%) | <0.001 |
| Anti-hypertensive | 0 (0.0%) | 7,371 (56%) | <0.001 |
| Anti-coagulant | 175 (0.4%) | 328 (3%) | <0.001 |
| Acetylsalicylic acid | 549 (1%) | 1,160 (9%) | <0.001 |
| Beta-blocker | 0 (0.0%) | 5,378 (41%) | <0.001 |
| Calcium antagonists | 0 (0.0%) | 1,983 (15%) | <0.001 |
| Diuretics | 0 (0.0%) | 4,689 (36%) | <0.001 |
| Statins | 1886 (4%) | 2,814 (21%) | <0.001 |
| Alternative cholesterol lowering medication | 95 (0.2%) | 119 (0.9%) | <0.001 |
| Anti-diabetics | 592 (1%) | 1,004 (8%) | <0.001 |
| **Health characteristics** |  |  |  |
| Diagnosed hypertension | 0 (0.0%) | 12,291 (93%) | <0.001 |
| Diagnosed hypercholesterolemia | 7,657 (16%) | 3,974 (30%) | <0.001 |
| Diagnosed diabetes | 1,142 (2%) | 1,428 (11%) | <0.001 |
| Systolic blood pressure | 136.00 [131.00, 143.00] | 135.00 [125.00, 146.00] | <0.001 |
| Diastolic blood pressure | 81.00 [75.00, 86.00] | 78.00 [72.00, 85.00] | <0.001 |
| Total cholesterol | 5.20 [4.60, 5.90] | 5.20 [4.50, 5.90] | <0.001 |
| LDL cholesterol | 3.44 (0.92) | 3.36 (0.93) | <0.001 |
| HDL cholesterol | 1.40 [1.20, 1.70] | 1.40 [1.10, 1.60] | <0.001 |
| Triglycerides | 1.11 [0.80, 1.58] | 1.26 [0.91, 1.76] | <0.001 |
| Renal function | 96.41 [86.10, 100.00] | 88.45 [76.94, 97.76] | <0.001 |
| *Data is presented as mean (SD), median [Q25, Q75] and n (%).* |  |  |  |

**Supplemental table 2.** Hazard ratios (HR) with 95% confidence intervals (95% CI) for the association between moderate to vigorous physical activity and stroke in medicated and non-medicated hypertensives.

| Total PA (MET min/week) | N sample | N outcomes | Unadjusted HR, P value | Model 1<br>HR (95% CI), P-value * | Model 2<br>HR (95% CI), P-value # |
| --- | --- | --- | --- | --- | --- |
| **Non-medicated hypertensives** |  |  |  |  |  |
| **Continuous** | 48,452 | 306 | 1.00 [0.999-1.00] | 1.00 [1.00-1.00] | 1.00 [1.00-1.00] |
| **P for linear trend** |  |  | 0.003 | 0.15 | 0.16 |
| **Quartiles** |  |  |  |  |  |
| Q1<1830 | 11,904 | 93 | REF | REF | REF |
| Q2 1830-3617 | 11,766 | 75 | 0.82 [0.60-1.11], *P* = 0.20 | 0.85 [0.61-1.19], *P* = 0.34 | 0.85 [0.61-1.18], *P* = 0.32 |
| Q3 3618-7175 | 12,197 | 74 | 0.78 [0.58-1.06], *P* = 0.12 | 0.67 [0.47-0.95], *P* = 0.02 | 0.67 [0.47-0.94], *P* = 0.02 |
| Q4 >7175 | 12,585 | 64 | 0.66 [0.48-0.91], *P* = 0.01 | 0.81 [0.57-1.16], *P* = 0.25 | 0.81 [0.57-1.16], *P* = 0.26 |
| **Medicated hypertensives** |  |  |  |  |  |
| **Continuous** | 13,169 | 161 | 1.00 [0.999-1.00] | 1.00 [1.00-1.00] | 1.00 [1.00-1.00] |
| **P for linear trend** |  |  | 0.58 | 0.89 | 0.85 |
| **Quartiles** |  |  |  |  |  |
| Q1<1830 | 3,916 | 52 | REF | REF | REF |
| Q2 1830-3617 | 3,329 | 36 | 0.82 [0.53-1.25], *P* = 0.35 | 0.94 [0.60-1.48], *P* = 0.80 | 0.96 [0.61-1.53], *P* = 0.89 |
| Q3 3618-7175 | 3,340 | 40 | 0.91 [0.60-1.37], *P* = 0.65 | 0.86 [0.54-1.36], *P* = 0.51 | 0.88 [0.56-1.39], *P* = 0.58 |
| Q4 >7175 | 2,584 | 33 | 0.97 [0.62-1.49], *P* = 0.88 | 1.12 [0.68-1.85], *P* = 0.64 | 1.15 [0.70-1.89], *P* = 0.59 |

